# Self-modulation of motor cortex activity after stroke alters behavior and corticospinal tract structure: a randomized controlled trial

**DOI:** 10.1101/2021.09.23.21263954

**Authors:** Zeena-Britt Sanders, Melanie K. Fleming, Tom Smejka, Marilien C. Marzolla, Catharina Zich, Sebastian W. Rieger, Michael Lührs, Rainer Goebel, Cassandra Sampaio-Baptista, Heidi Johansen-Berg

## Abstract

Real-time functional magnetic resonance imaging (fMRI) neurofeedback allows individuals to self-modulate their ongoing brain activity. This may be a useful tool in clinical disorders which are associated with altered brain activity patterns. Motor impairment after stroke has previously been associated with decreased laterality of motor cortex activity. Here we examined whether chronic stroke survivors were able to use real-time fMRI neurofeedback to increase laterality of motor cortex activity and assessed effects on motor performance and on brain structure and function. We carried out a randomized, double-blind, sham-controlled trial in which 24 chronic stroke survivors with mild to moderate upper limb impairment experienced three training days of either Real (n=12) or Sham (n=12) neurofeedback. Stroke survivors were able to use Real neurofeedback to increase laterality of motor cortex activity within, but not across, training days. Improvement in gross hand motor performance assessed by the Jebsen Taylor Test (JTT) was observed in the Real neurofeedback group compared to Sham. However, there were no improvements on the Action Research Arm Test (ARAT) or the Upper Extremity Fugl-Meyer (UE-FM) score. Additionally, decreased white-matter asymmetry of the corticospinal tracts was detected 1-week after neurofeedback training, indicating that the tracts become more similar with Real neurofeedback. Changes in the affected corticospinal tract was positively correlated with neurofeedback performance. Therefore, here we demonstrate that chronic stroke survivors are able to use fMRI neurofeedback to self-modulate motor cortex activity, and that training is associated with improvements in hand motor performance and with white matter structural changes.

## INTRODUCTION

Stroke survivors often suffer debilitating long-term motor impairments of the upper-limb, which can have a negative impact on activities of daily living *(1)* and quality of life *(2)*. Rehabilitation typically focuses on the stroke affected limb, however, there has been increased interest in interventions that target the brain (e.g. brain stimulation), in the hope of boosting rehabilitation effects and further improving motor function. These studies typically target motor cortex activity and many attempt to promote lateralization of activity towards the stroke-affected motor cortex (for review: *3, 4)*.

A novel alternative approach, which harnesses the brains intrinsic capacity for activity modulation, is for stroke survivors to learn to self-modulate brain activity through real-time fMRI neurofeedback (NF). Previous work has shown some promising behavioral effects of fMRI NF (e.g. *5–7)*, and fMRI NF has been explored in various clinical conditions associated with aberrant brain activity patterns, for example depression *(8, 9)*, pain disorders *(10, 11)* and phobias *(12, 13)*. FMRI NF has also been suggested as a potentially useful tool in stroke rehabilitation *(14, 15)*, however to date no randomized, sham-controlled trials have been published. Previous pilot studies have shown that stroke survivors appear to be able to use fMRI NF to control activity in a variety of motor regions *(16–18)* demonstrating feasibility of this approach. After sufficient training, participants receiving NF training may be able to maintain the ability to self-modulate brain activity outside of the training sessions, leading to potentially long-lasting improvements (for review see: *(19)*).

Here we carried out a registered double-blind, sham-controlled trial to investigate the efficacy of three sessions of real-time fMRI NF in chronic stroke survivors with ongoing upper-limb impairments. The primary outcome measures of this trial were changes in *lateralization of motor cortex activity* during movements of the stroke affected hand, and changes in *motor performance* as assessed using the Jebsen Taylor Hand Function Test (JTT; *(20)*). Additionally, secondary outcomes included measures of short-term and long-term NF learning transfer, where participants’ ability to maintain changes in brain activity was assessed, as well as changes in clinical scores and brain structure.

## RESULTS

A double-blind, sham-controlled, between-subjects study was carried out to investigate chronic stroke survivors’ ability to use fMRI NF, and its effects on behavior and brain structure and function. One hundred and seventy-three chronic stroke survivors (>6 months post stroke) were contacted and assessed for eligibility (see recruitment flow chart **Supplementary figure S1**), of whom 24 were randomized into the Real (n=12) or the Sham (n=12) NF groups. Randomization included minimization of variance in baseline score on the Action Research Arm Test (ARAT) *(21)* and time since stroke. Baseline participant characteristics are displayed in *table 1*.

**Table 1:** Participant demographics for the Real (top) and Sham (bottom) group. Group median and range indicated where appropriate. F, female; M, male; L, left; R, right; Note: Fugl Meyer Upper Extremity assessment (UE-FM) was scored by two independent blinded assessors and the average score was taken.

|  | Number of<br>participants | Age<br>(years) | Sex | Time post-stroke<br>(months) | Baseline<br>ARAT | Baseline<br>UE-FM | Hand<br>affected |
| --- | --- | --- | --- | --- | --- | --- | --- |
| **Real**<br>**group** | 12 | 63<br>(50-84) | 4F/8M | 54.5<br>(11-230) | 36.5<br>(8-54) | 44<br>(28.5-60) | 2R/10L |
| **Sham**<br>**group** | 12 | 59.3<br>(36-86) | 1F/11M | 62.5<br>(7-237) | 35<br>(12-55) | 44.5<br>(18.5-63) | 6R/6L |

Participants were invited to attend six testing days in total (see Figure 1A). Measures of upper limb function and impairment (ARAT, upper extremity Fugl-Meyer assessment (UE-FM), JTT) and neuroimaging measures (MRI and electroencephalogram; EEG) were collected at Baseline and 1-week follow-up. The motor assessments only were also repeated at a 1-month follow-up. Participants underwent three NF training days, with the first two training days separated by 24 hours and the second and third training days by 48 hours. The JTT and EEG measures were also carried out at the end of each NF training day.

**Figure 1:**
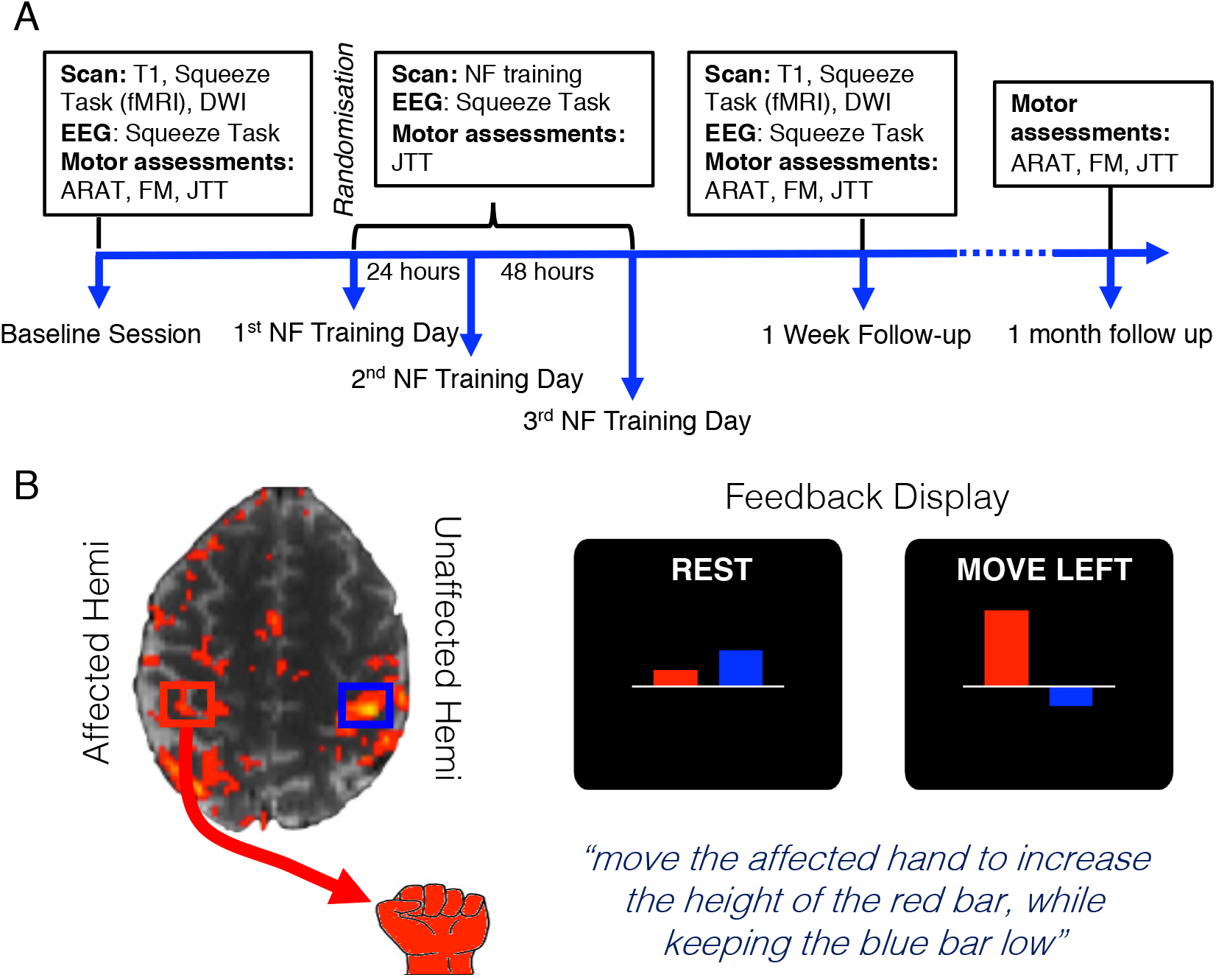
Study design and timeline. **(A)** Participants attended three NF training sessions separated by 24 and 48 hours, a baseline session and two follow-up sessions at 1-week and 1-month post NF training. **(B) Left:** Regions of interest (ROIs) used for NF training were defined during a functional localizer. ROIs (red and blue squares) were centered on peak activity in M1 during movements of both hands. **Right**: During NF training, only the stroke-affected hand was moved and participants viewed two bars on the screen. The red bar represented activity in the stroke affected hemisphere and the blue bar represented activity in the unaffected hemisphere. During movement blocks, participants were instructed to make movements with their stroke affected hand to increase the size of the red bar, while keeping the blue bar as low as possible.

During NF training, participants observed two moving vertical bars whose heights represented ongoing activity in the stroke affected (red bar) and unaffected (blue bar) primary motor cortices (Figure 1B). Participants were instructed to move their stroke-affected hand and try to increase the height of the red bar, while keeping the blue bar low. This corresponded to increasing laterality of brain activity in the primary motor cortices (M1) towards the stroke-affected hemisphere. During each training day, participants completed three runs of NF (or Sham) training (see **Materials and Methods: Neurofeedback Intervention**). On each day, before and after NF training, NF learning transfer was assessed by asking participants to move their stroke-affected hand while attempting to regulate their brain activity in the absence of NF. A debriefing questionnaire was completed after every training day, which showed that both groups experienced similar control over the bars (**Supplementary figure S2**).

### FMRI during NF training days (Figure 2)

#### Primary outcome: Laterality of M1 activity increases with Real NF within days but not across days

The first primary outcome measure was the change in laterality of M1 activity during NF training. This was assessed first *within* training days and then *across* training days. A laterality index (LI) of brain activity in the Regions-of-interests (ROIs) used for the NF training was calculated (see **Materials and Methods: Laterality and Success Index Calculation**) for each NF run on each NF training day (3 runs per day, 9 in total; see Figure 2A). Baseline LI was calculated as the LI during the first transfer run and was included as a covariate in statistical analysis to correct for any baseline differences in LI.

**Figure 2:**
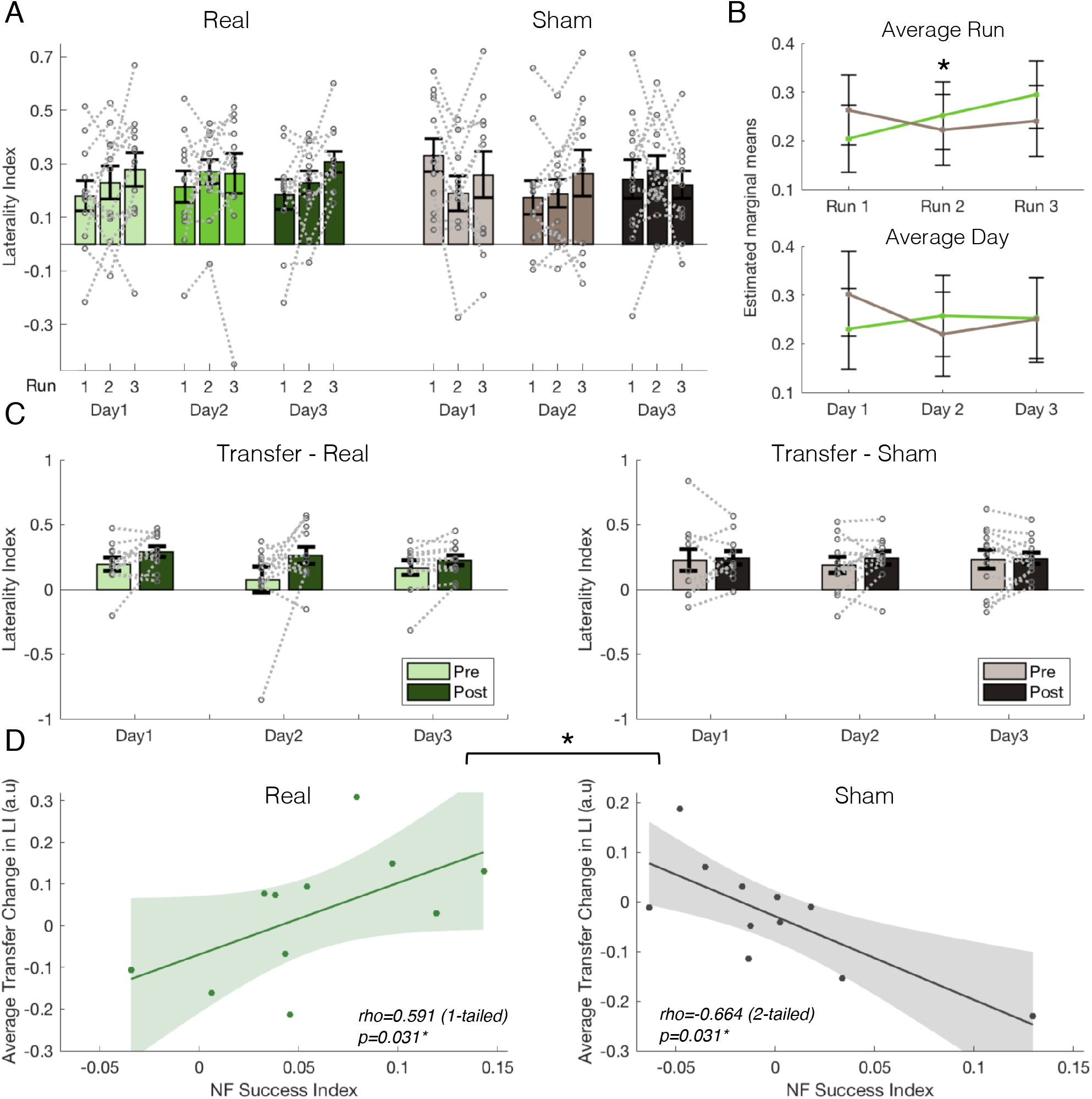
NF learning occurs within training days and better learning is associated with transfer success. **(A)** Laterality of M1 activity (LI) is displayed for the Real (green/left, n=12) and Sham (grey/right, n=12) group over all the training runs and days. Bars represent group means, grey lines show individual participant data and error bars represent s.e.m. **(B) Top**: When considering NF learning *within* training days by averaging LI for each run (pooled over training days), a significant interaction could be observed between Run and Group *(p*=0.037), with the Real group increasing LI over runs, whilst the Sham group didn’t change. Estimated marginal means and confidence intervals are shown for each run. **Bottom**: When assessing NF learning *across* training days by averaging LI for each day (pooled over runs), there were no significant interaction or main effects. **(C)** There was no group-level transfer effect; participants in both groups have similar change in LI on the transfer runs (data displayed as in A). **(D)** However, the relationship between transfer change and NF success differs between the groups (p=0.002), with a positive relationship found for the Real group, and a negative one for the Sham group. Shaded areas represent 95% confidence intervals. *p<0.05

In order to assess NF learning *within* training days, LI for each of the 3 runs was averaged across all training days for each participant, resulting in average LI values for the first, second and third run, pooled across training days (Figure 2B top). A Linear Mixed Model (LMM) was fit to the average run LI values, with fixed effects of Group and Run and their interaction, Baseline LI as a covariate, and random intercepts per participants in the random effects structure. A significant Group×Run interaction was found (F(2,46)=3.556, p=0.037). Post-hoc pairwise comparisons (using Tukey correction for multiple comparisons) showed that this interaction was driven by a significant increase in LI between Run 1 and Run 3 in the Real group (t(50.4)=2.815, p=0.019, Run1-Run3: *b*=-0.090, SE=0.032) (Figure 2A and 2B). No significant differences between runs were found in the Sham group (all: t(50.4)<1.5, p>0.1). This indicates that the Real NF group were able to increase the laterality of their M1 activity, towards the affected hemisphere, over the runs. Exploration of changes in activity in each hemisphere separately revealed no significant group effect or interaction (**Supplementary table 1**). A voxelwise analysis examining differences between average run activity in the whole brain, revealed increased activity in the putamen of the unaffected hemisphere in the Real NF group during the first NF runs before decreasing (see **Supplementary figure S3**).

Next NF learning *across* training days was assessed by averaging LI over runs within each training day for each participant, resulting in average LI values for each day (Figure 2B bottom). A LMM was fit to the average day LI values in the same way as described above, but with the fixed effect of Day rather than Run. There was no significant Group×Day interaction (F(2,46)=1.061, p=0.355), or main effect of Group (F(1,23)=0.065, p=0.800), or Day (F(2,46)=0.262, p=0.771), suggesting that LI scores did not change over days and that day to day changes in LI did not differ across groups (Figure 2A and 2B).

#### Secondary outcomes: No evidence for short-term transfer effects following NF training

Secondary fMRI outcomes included exploring participants’ ability to maintain NF learning when feedback was removed. This was assessed during transfer runs before and after NF training on each day. Participants were instructed to use the strategies they felt were most successful but were not shown the NF bars. Again, laterality of brain activity during the transfer runs was assessed by calculating a LI based on activity in each of the ROIs. To assess whether there was a within-day transfer effect, a change score was calculated between pre- and post-transfer LI on each day. A LMM was carried out on this data with fixed effects of Group and Day and their interaction, and random intercepts in the random effects structure. There was no significant interaction (F(2,44)=0.417, p=0.662) or main effects (Group: F(1,23)=1.642, p=0.213; Day: F(2,44)=2.276, p=0.115) (Figure 2C). This suggests that changes in laterality of M1 activity with NF did not persist after the feedback was removed.

#### Secondary outcomes: Performance in short-term transfer test is correlated with NF success

High LI variability during NF training and in the transfer runs was observed, with some participants showing large increases in LI, and others showing little or no change in LI (Figure 2A&C). It was hypothesized that participants who were more successful at using NF would also show the largest increase in LI on the transfer task. In order to quantify how successful individual participants were at using NF, a “NF success index” was calculated as a regression slope over LI of NF runs for each day, and then averaged over training days (see **Materials and methods: Laterality and Success Index Calculation**). Where values were missing (see **Supplementary methods: Missing data**), the slope across the remaining runs was taken.

Average transfer change was calculated as the post-pre difference in LI on the transfer runs per participant (adjusted for baseline LI) averaged across the three training days. The predicted positive correlation was observed in the Real group (Figure 2D), (n=11, Spearman’s rho=0.591, p=0.031, one-tailed), whereas a negative correlation was observed in the Sham group (n=11, rho=-0.664, p=0.031, two-tailed). A Fisher’s Z-test confirmed that the two correlations were significantly different from each other (Z=2.958, p=0.002) (Figure 2D).

### Behavioral and clinical measures (Figure 3)

#### Primary outcome: Gross motor hand performance improves following Real NF training

The second primary outcome measure of this trial was motor performance changes on the JTT. Performance on the JTT was measured at baseline, after each NF (or sham) training session, 1-week after training and 1-month later. A LMM was carried out on the average time (log-transformed) over all sub-tasks at each time point, with Group and Day as fixed factors, baseline time as a covariate and random intercepts in the random effects structure. This revealed a main effect of Time (F(4,90.326)=3.207, p=0.017), with participants in both groups improving over time (post-hoc linear contrast: t(99.3)=-2.818, p=0.006, *b*=-0.205, SE=0.073). However, there was no main effect of Group (F(1,24.225)=2.660, p=0.116) or Group×Time interaction (F(4,90.326)=1.388, p=0.2445) (Figure 3A).

**Figure 3:**
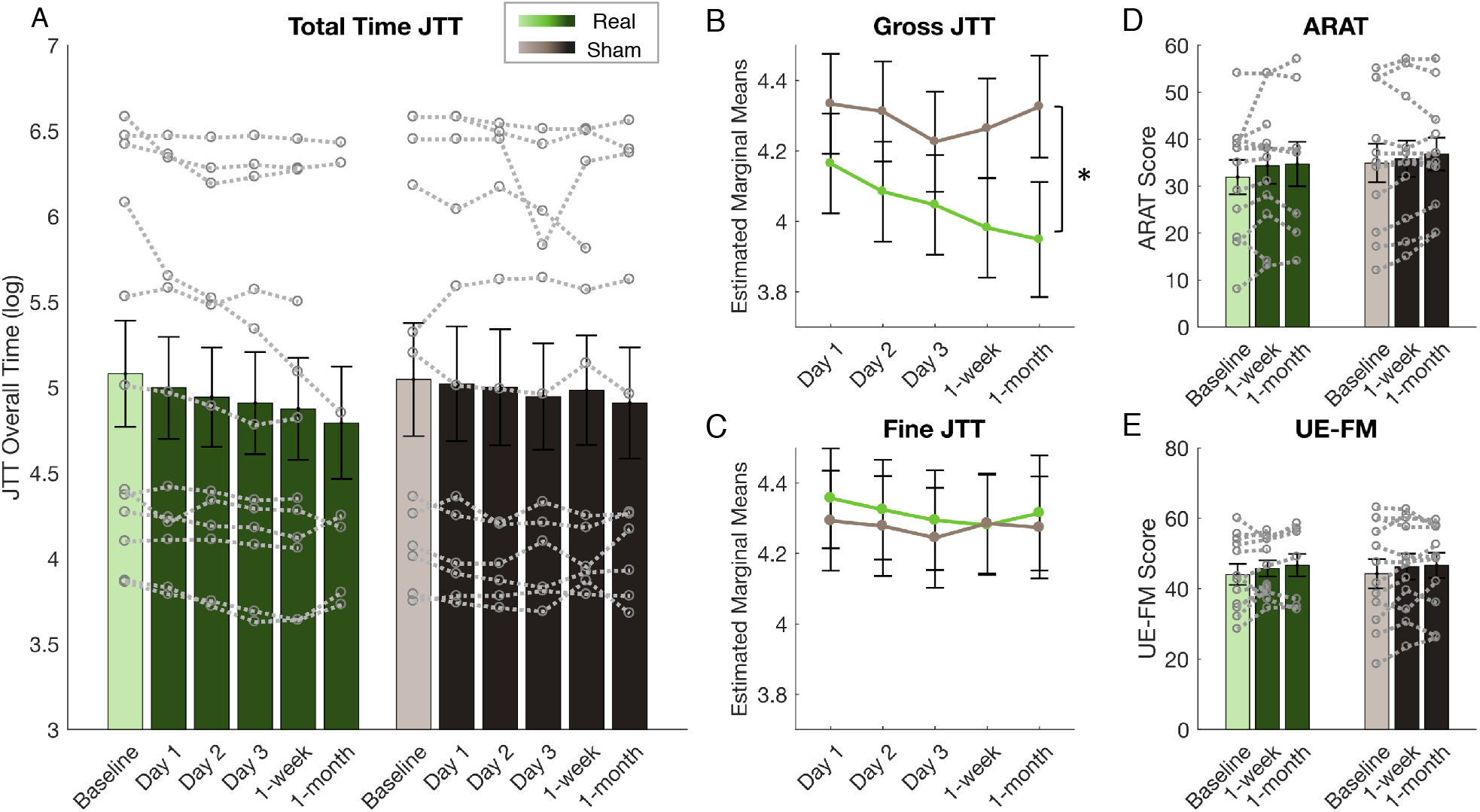
Participants in Real group improve on Gross motor performance (JTT) compared to Sham. **(A)** Participants in both the Real (green) and the Sham (grey) group got faster at performing the JTT over time when all subtasks were considered (p=0.017) and no group differences were found. Bars represent mean time to complete all subtasks (log), error bars represent s.e.m and data from individual participants is shown by the grey dotted lines. (**B)** When comparing performance on the Gross motor sub-tasks of the JTT, the Real group had faster performance compared to the Sham group (p=0.01). Estimated marginal means are shown with 95% confidence intervals at each follow-up timepoint. * p<0.05. (**C)** The same data is shown for Fine motor sub-tasks of the JTT where no significant group differences were found. There was no group effect on the ARAT (**D)** or the UE-FM (**E**) at 1-week or 1-month after NF training. Data displayed as in (A).

In line with previous research *(22–24)*, we also separately analysed performance on Gross motor sub-tasks of the JTT (stacking checkers, moving heavy objects, moving light objects) and Fine motor sub-tasks of the JTT (card turning, picking up small objects, simulated feeding). A LMM was carried out with Group, Day and Task (Gross, Fine) as fixed effects, baseline performance as a covariate and random intercepts in the random effects structure. There was a significant main effect of Task (F(1,168)=33.374, p<0.001) and Group×Task interaction (F(1,183.32)=42.05, p<0.001). Post-hoc pairwise comparisons revealed that this effect was driven by faster performance in the Real group compared to the Sham group on the Gross motor tasks (t(34.8)=-2.711, p=0.01; Real–Sham: *b*=-0.247, SE=0.091) (Figure 3B). In contrast, there was not a significant difference between the two groups on performance of the Fine motor tasks (Figure 3C; t(34.8)=0.432, p=0.668; Real-Sham: *b*=0.039, SE=0.091). Whilst the main effect of Day approached significance (F(4,156.05)=2.412, p=0.051), there were no other significant main effects of interactions (all F<2, p>0.1). Overall, these findings suggest that participants in the Real group had improved performance on gross motor tasks compared to the Sham group.

#### Secondary outcomes: No evidence for Real NF effects on the ARAT or UE-FM

Secondary outcome measures included changes in participants’ performance on two clinical measures, the ARAT and the UE-FM (Figure 3D&E). Performance on these measures was assessed at baseline, 1-week post intervention and 1-month later. When comparing the two groups at the 1-week follow-up, with baseline as a covariate, the two groups were not significantly different from each other on the ARAT (F(1,21)=0.717, p=0.407, 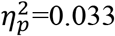), or the UE-FM (F(1,21)=0.057, p=0.813, 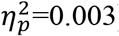). Similarly, at the 1-month follow-up there was also no difference between the groups on either measure (ARAT: F(1,15)=0.308, p=0.587, 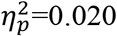. UE-FM: F(1,15)=0.055, p=0.818, 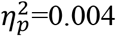). When all time points were considered in a LMM, both groups improved over time (see **Supplementary materials: Improvements over time on ARAT and UE-FM**).

### White matter structure (Figure 4)

#### Secondary outcome: Alterations in the corticospinal tract structure after Real NF training

Before and 1-week after NF training, diffusion-weighted imaging (DWI) was acquired to assess microstructural changes in participants’ white matter. Fractional anisotropy (FA) was extracted from the stroke affected and unaffected corticospinal tracts (CST) and FA-asymmetry was calculated (see **Materials and Methods: MRI analysis**). FA asymmetry of the CSTs has previously been linked with stroke impairment *(25, 26)* and was therefore used as the measure of interest here. FA asymmetry decreased (between baseline and 1-week) in the Real NF group and increased in the Sham group (Figure 4A). An ANCOVA comparing 1-week follow-up values between the two groups, with baseline FA asymmetry as a covariate, revealed a significant group difference (F(1,18)=8.99, p=0.008; 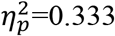), with the Real group having significantly lower FA asymmetry after NF training than the Sham group (EMM: Real=0.089, Sham=0.098) when accounting for baseline.

**Figure 4:**
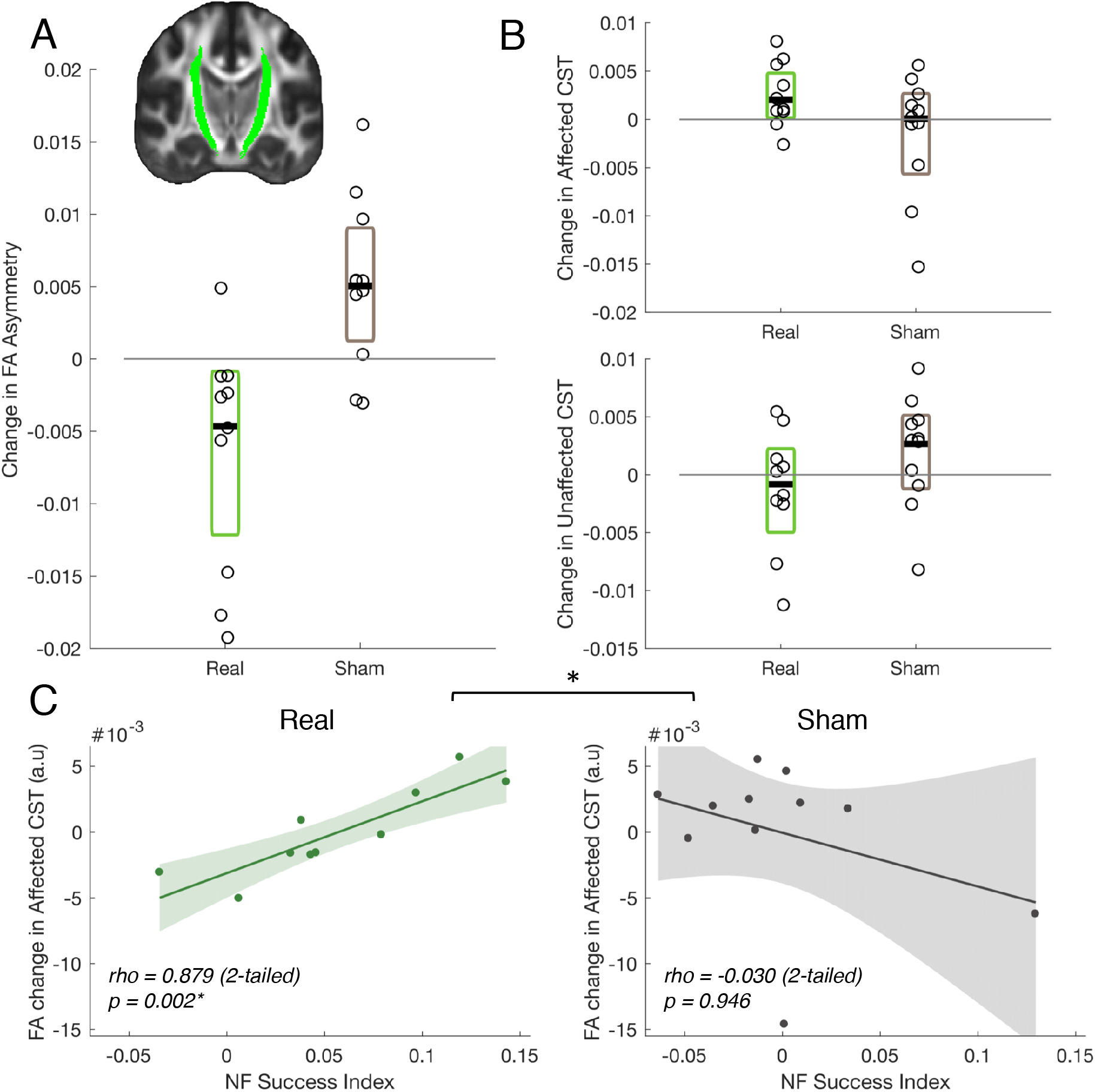
FA Asymmetry of CSTs is reduced after Real NF training. **(A)** The Real group (n=10) had lower FA asymmetry in the CSTs (shown in inset) after NF training compared to the Sham group (n=11). Change in FA asymmetry between baseline and 1-week follow-up is plotted for the Real (green) and the Sham (grey) group. The black line represents median change, colored boxes represent 95% confidence intervals and individual participant data points are shown with open circles. **(B)** The same information is shown for FA change between baseline and 1-week follow-up in the affected (top) and unaffected (bottom) CSTs. **(C)** Neurofeedback success was positively correlated with FA change in the stroke affected hemisphere in the Real group, whereas no correlation was found in the Sham group with the two correlations being significantly different from each other *(p* = 0.004). * p<0.05

Effects on FA asymmetry can be driven by one or both hemispheres; changes in FA for each hemisphere separately are shown in Figure 4B. Follow-up ANCOVAs comparing the two groups on each CST separately failed to reach significance (Group effect Affected Hemi: F(1,18)=2.501, p=0.131, 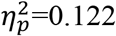; Unaffected Hemi: F(1,18)=1.442, p=0.245, 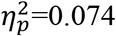), but exploratory pairwise comparisons suggest that the change in asymmetry was more likely driven by increases in FA in the Affected CST in the Real group (t(9)=-2.338, p=0.044, *d*=-0.739), rather than changes in the Unaffected CST (t(9)=0.841, p=0.422, *d*=0.266), or the Sham group (Affected: t(10)=0.795, p=0.455, *d*=0.240; Unaffected: t(10)=-1.380, p=0.198, *d*=-0.416). However, the pairwise difference for the Affected CST in the Real group would not survive correction for multiple comparisons and should be interpreted with caution.

#### FA change in the affected corticospinal tract is correlated with NF training success

Additionally, the relationship between FA changes and NF success was also explored. NF success did not correlate with changes in FA asymmetry (adjusted for baseline asymmetry) in the Real group (Spearman’s rho=-0.297, p=0.407), or the Sham group (Spearman’s rho=-0.164, p=0.657). However, there was a significant correlation between change in FA in the Affected CST (adjusted for baseline FA; Figure 4C) and NF success in the Real group (Spearman’s rho=0.879, p=0.002; Bonferroni corrected alpha for 6 comparisons=0.008), but not in the Sham group (Spearman’s rho =-0.030, p=0.946). A Fisher’s Z-test confirmed the two correlations were significantly different from each other (Z=2.622, p=0.004). In contrast, correlations with change in FA in the Unaffected CST did not reach significance in either group (Real: rho=0.612, p=0.067, Sham: rho=-0.297, p=0.407). This suggests that participants who were more successful at using NF also had the greatest increases in FA in the stroke-affected CST.

### Transfer to visuomotor squeeze task (Figure 5)

#### Secondary outcome: No transfer to visuomotor squeeze task

Finally, we used a visuomotor squeeze task to test for any transfer of activity modulation with NF to a constrained task (i.e. same movement parameters pre-post). Participants were instructed to squeeze on a force transducer with their stroke-affected hand at a specific rate and force (see **Materials and methods: Visuomotor squeeze task**) therefore this transfer task differs from the task used during NF training where participants could move their affected hand in any way they wished. Brain activity during the squeeze task was assessed using fMRI and EEG on the baseline and follow-up days, as well as after each NF session using EEG.

**Figure 5:**
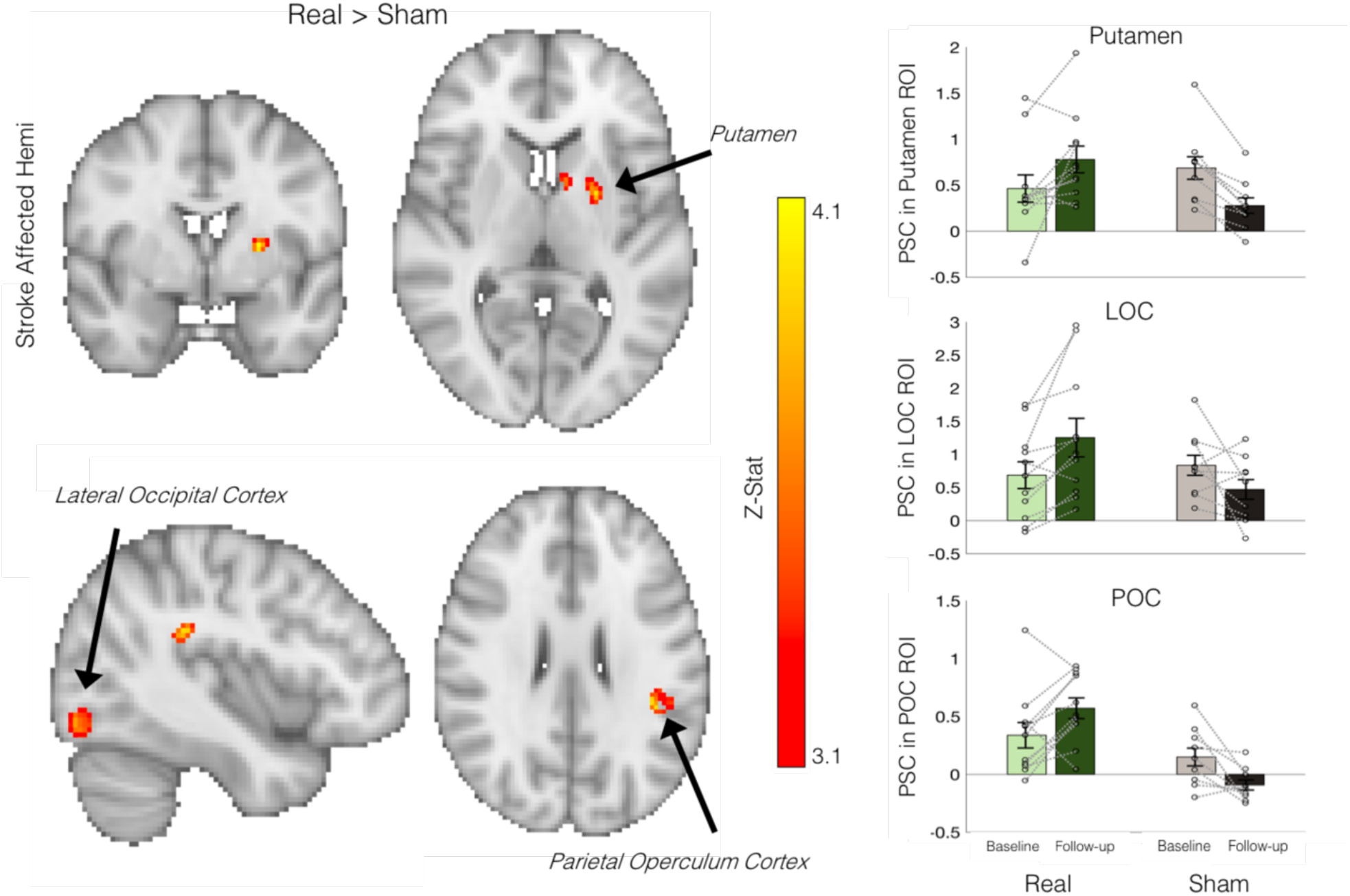
Increased activity in brain regions associated with NF learning. Changes in brain activity before and after NF or Sham training were assessed during a controlled visuomotor squeeze task. Three significant clusters were found where the Real group had greater change in activity following NF than the Sham group (Real>Sham, voxelwise GLM, p<0.05, corrected), located in the putamen, lateral occipital cortex (LOC) and the parietal operculum cortex (POC) of the unaffected hemisphere. For visualization purposes, the mean percent signal change (PSC) of the significant clusters is plotted on the right, as well as the data from individual participants (represented by open circles). Error bars represent s.e.m.

For the fMRI task, a LI was calculated based on activity in an anatomically defined hand-knob ROI. A between-subjects ANCOVA on follow-up LI in the two groups, with baseline LI as a covariate, showed no significant differences between the two groups (F(1,17)=0.166, p=0.689, 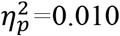). LI at follow-up (adjusted for baseline LI) in the Real group was also not correlated with the NF success index (rho=-0.009, p=0.989, n=11, two-tailed). Additionally, no evidence of transfer effects were observed on the EEG visuomotor squeeze task where LI was calculated over all sessions based on the event related desynchronization (ERD) obtained from electrodes C3 and C4 (sensorimotor cortex; see **Supplementary figure S4**).

#### Increased activity in NF learning related brain regions after training

Whole-brain voxelwise analysis comparing the difference between follow-up and baseline brain activity, across the two groups (Real vs Sham) revealed three significant clusters (Figure 5). Significant clusters were all located in the unaffected hemisphere and included the putamen (p=0.015, MNI coordinate of max T-stat: −24 0 8), the lateral occipital cortex (LOC; p=0.041, - 38 −82 −14) and the parietal operculum cortex (POC; p=0.047, −42 −38 26). In each cluster the Real group increased activity after NF training, whereas the Sham group decreased or stayed the same. Finally, within the Real group we tested whether baseline brain activity during the squeeze task is associated with subsequent NF performance. Whilst there was no association between laterality of M1 activity at baseline and NF success index (rho=-0.336, p=0.290, two-tailed), two significant clusters were found when exploring associations across the whole brain (see **Supplementary figure S5**).

## DISCUSSION

Our study provides the first evidence from a randomized, double-blind, sham-controlled trial that chronic stroke survivors are able to use fMRI NF to increase laterality of M1 activity during motor execution. Further, NF training was associated with improvements in gross hand motor performance and alterations in white matter microstructure. This study therefore provides important new evidence on opportunities and challenges for translation of fMRI NF to the setting of stroke rehabilitation.

One key challenge for any clinical NF intervention is to deliver performance gains that persist after NF is removed. Comparing Real to Sham NF, we did not find NF carry-over effects across training days, nor transfer effects when the NF display was removed. However, consistent with prior literature *(27, 28)*, NF training success was highly variable across participants in the Real NF group. Importantly, this variability in NF success during training was linked to the persistence of NF gains. Participants who were more successful at self-modulating brain activity during NF, were also more likely to be able to increase motor activity laterality in the absence of NF. However, there was no evidence of this association on a constrained visuomotor squeeze task, suggesting that this association only applies to certain movements. Research has begun to investigate factors related to variability in NF learning success (e.g. *28–30)*, which could allow future studies with larger samples to identify individuals most likely to respond to NF training using baseline measures.

Another key challenge for the development of clinical NF interventions is to drive changes in brain activity that translate into meaningful and long-lasting improvements in clinical symptoms. Here we tested motor function using a range of measures and found improved performance on the gross motor sub-tasks of the JTT in the Real group compared to the Sham group. That improvements were found for gross rather than fine sub-tasks fits with extensive evidence that fine motor tasks are the hardest to recover following damage to the corticospinal system *(31–33)*.

We did not find any evidence of NF training improving scores on clinical scales of impairment (UE-FM) or activity (ARAT). This lack of an effect on clinical measures may be due to a number of factors. One possibility is that laterality of brain activity may not be a suitable intervention target for all stroke survivors. In particular, it has previously been suggested that more severely affected stroke survivors may instead benefit from boosting activity in both M1’s *(34)*. Additionally, neuromodulation approaches are often considered not as treatments per se, but rather as approaches that enhance the potential for use-dependent plasticity *(35)*. Therefore, the addition of further motor rehabilitation outside of the scanner, to drive the motor system while it is ‘primed’, may be necessary to elicit observable effects of NF training on clinical measures. For example, a previous study in Parkinson’s patients found clinically relevant improvements of symptoms after just two sessions of NF combined with extensive practice outside of the scanner using the successful strategies identified during training *(36)*. However, the finding of an effect on the gross sub-tasks of the JTT is a promising first indication that NF can induce behavioral change in this population, and ways to bolster this effect should be explored further. Especially considering that the current study NF training was conducted on a relatively short timescale (3 sessions of 20mins) and without further practice outside the scanner. Previous studies which demonstrated significant motor improvements in chronic stroke survivors are typically conducted over a much longer time period consisting of many hours of training over multiple days or weeks (for example ∼9 hours using TDCS: *(37)*, ∼7 hours of EEG NF: *(38)*, or between ∼17 and ∼90 hours of motor training *(39, 40)*).

Any long-term benefits of NF are likely to be mediated via structural and functional brain plasticity in brain systems engaged in training. To test for structural plasticity, we assessed FA asymmetry in the CSTs, as this measure is consistently found to correlate with motor dysfunction after stroke *(41)*, therefore finding ways to strengthen (or rewire) CST output could potentially lead to improved functioning *(42)*. NF training led to reduced FA asymmetry and participants who performed better on the NF training exhibited greater increases in CST FA of the affected hemisphere. These results provide the first evidence in a clinical population that fMRI NF can induce structural changes in white matter tracts and support previous work showing similar effects in healthy individuals targeting motor areas *(43, 44)*. Recent rodent studies have shown that optogenetically activating the intact contralesional CST after stroke, in combination with intense rehabilitation, led to near-complete recovery of skilled forelimb function as well as corticospinal sprouting from the intact CST to the denervated tract *(45)*. In humans, whilst white matter plasticity plays an important role in learning *(46)*, there is little evidence of neuromodulation techniques that directly target or boost white matter plasticity. Recent work using TMS NF has demonstrated the feasibility of directly modulating excitability of CSTs in healthy participants *(47)*, however, it remains to be discovered whether this can lead to behavioral improvements in healthy people or stroke survivors. Our results show that fMRI NF might be a promising tool to target CST structure in stroke survivors, potentially leading to better motor recovery.

In addition to changes in brain structure after NF, we also found functional brain changes during a visuomotor squeeze task. After Real NF training, increased activity in several brain regions which have been implicated in NF learning was observed, including the putamen (dorsal striatum), LOC and the POC. Currently, the precise mechanism underlying NF learning remains unknown, however the dorsal striatum has been consistently implicated in NF learning *(29, 48–50)*; and blocking long term potentiation in the striatum impairs NF learning *(51)*. This has led to the suggestion that NF learning may involve procedural and reinforcement learning *(49, 50)*. Here we also demonstrated increases in putamen activity during initial NF learning (**Supplementary figure S3**). The finding that activity in the putamen is also increased 1-week after NF training suggests that participants in the Real group may have learnt to engage this area during initial motor performance. Previous research in rodents has shown that connectivity between the dorsal striatum and motor cortex increases with NF training (33,34), which may lead to increased activity in the dorsal striatum when the motor cortex is engaged.

The LOC has been demonstrated to be a brain region consistently activated during NF learning irrespective of the NF target *(48)*. The LOC is involved in directing attention to visual signals *(53)*, and may reflect participants in the Real group learning to pay more attention to the visual display after NF training. Additionally we found increased activity in the POC, where the secondary somatosensory cortex (SII) is located. SII has previously been associated with tactile learning *(54)* and decision making *(55)*. The identification of brain regions that are involved in NF learning helps to improve understanding of mechanisms involved in NF learning, which could help to optimize future studies and could also have implications for individuals who may have damage to these regions.

There were several limitations to this trial. Whilst the sample size in the current study was favorable compared to previous neuromodulation studies in this population, replication is needed in a larger sample of stroke survivors, that would allow more exploration of variability in response. Furthermore, the addition of motor rehabilitation and more training sessions could establish whether clinically relevant changes in behavior can be achieved with fMRI NF. Using fMRI has the advantage of whole brain coverage, allowing to monitor changes throughout the brain which may shed light on mechanisms involved and off-target changes that may underlie changes in behavior. However, cheaper and more practical modalities such as EEG would lend themselves to the clinical translation of this approach. FMRI NF is a highly flexible approach and it has become increasingly transparent that it is unlikely that there is a one-size-fits-all rehabilitation solution for all stroke survivors, and the need to stratify individuals has been highlighted *(56, 57)*. Future research could harness the flexibility of this approach to personalize treatments to better suit individual stroke survivors.

## MATERIALS AND METHODS

### Study design

A double-blind RCT (ClinicalTrials.gov: NCT03775915) was carried out to examine chronic stroke survivors’ ability to use real-time fMRI NF to increase laterality of M1 activity during affected hand movements. Additionally, effects of NF on motor performance and brain structure and function were also assessed. A parallel design was selected to avoiding carry over effects, as it is currently unclear how long NF effects last.

One-hundred and seventy-three chronic (>6 months post-stroke) stroke survivors were contacted to take part in the study between January 2018 and March 2020. Of these, 118 stroke survivors were screened to assess whether they met the inclusion criteria (see recruitment flow chart **Supplementary figure S1**). Inclusion criteria included prior symptomatic stroke with ongoing effects on upper limb movements on one side of the body and some residual movement in the stroke-affected hand. Exclusion criteria included MRI scanning contraindications, previous history of neurological or psychiatric illness, and limited communication or inadequate understanding of instructions. The trial was terminated in March 2020 due to the COVID-19 lockdown before the full sample (30 participants) could be collected. Sample size was based on previous NF studies using similar study design *(44, 58)*.

Twenty-seven (22 males) chronic stroke survivors (>6 months post stroke), met the initial screening criteria and were recruited into the trial. All participants provided written informed consent in accordance with the Declaration of Helsinki and the National Research Ethics service (Oxford University, UK) approved protocol (14/LO/0020). Two participants withdrew from the trial after the initial baseline session and were therefore not randomized (one due to M1 lesion, one did not tolerate the MRI environment). One participant’s study participation was halted after the baseline session due to restrictions related to the COVID-19 pandemic. Twenty-four participants (19 males) were randomized following the baseline session (Real group=12, Sham group=12) and completed the NF training. Participants were randomized with minimization of variance in time-since-stroke and baseline ARAT score using a freely available randomization service (www.rando.la). However, the first three participants were forced into the Real group in order to ensure that sham videos were available. Participants were blind to which condition they had been allocated to throughout the experiment. Randomization was performed by a researcher (Z.B.S), who also set up the NF software to deliver the correct condition, but was not involved in any clinical assessments. All clinical assessments were carried out by blinded researchers (T.S, M.M, M.F). Blinded researchers also gave all task instructions during NF training and completed the debriefing questionnaires.

Participants were invited to attend six testing sessions which took place at the University of Oxford. During an initial baseline session, assessments of upper limb function were carried out, as well as baseline structural and functional neuroimaging measurements. After the baseline testing day, participants were randomized and attended three NF training sessions, during which they received either Real or Sham NF (see **Neurofeedback Intervention**). Participants then attended two follow-up testing days: one at 1-week post NF where all of the baseline measures were repeated, and another 1-month later where only the assessments of upper limb function were completed.

### Neurofeedback Intervention

NF training was carried out over three days, with 24 hours between the first two training days, and 48 hours between the second and third training days (Figure 1A). During each NF training day, a functional localizer was initially carried out, during which participants were instructed to open and close their hands at a steady rate that was comfortable for them. There were three 15 seconds movement blocks for each hand (6 in total), interspersed with 15 seconds rest blocks. Participants saw the instructions ‘Squeeze Right’, ‘Squeeze Left’ or ‘Rest’ on the screen, displayed using TurboFeedback (v1.0). A real-time general linear model (GLM) analysis was carried out using Turbo-BrainVoyager (v3.2, Brain Innovation B.V., Maastricht, The Netherlands) and used to identify the peak activity in the hand knob region of the sensorimotor cortex, and two ROIs (1.6×1.6×1cm) were centered on the peak activity in each hemisphere (Figure 1B).

Participants experienced three NF training runs on each day. During NF training runs participants saw two bars on the screen (Figure 1B), the height of which was determined by the measured percent signal change (PSC) in the ROIs (see **Supplementary methods: Online analysis**). Participants were instructed to try different movements with their stroke affected hand to increase the size of the red bar, whilst keeping the blue bar as low as possible. This corresponded to increasing activity in the ROI of the stroke affected hemisphere (represented by the red bar), while keeping activity in the ROI of the unaffected hemisphere to a minimum (represented by the blue bar). The height of the bars was continuously updated with each repetition time (TR).

Participants were given some suggested strategies that they could try, including opening and closing their stroke-affected hand or tapping individual fingers (see **Supplementary methods: Suggested strategies**), however it was made clear that they could use whatever strategies they wished. Each NF training run consisted of six 30-second NF blocks during which the instruction “Move Left” or “Move Right” (depending on which hand was stroke-affected), was displayed above the bars, and seven interspersed 30-second rest blocks, during which the instruction changed to “Rest”. Participants were explicitly instructed not to move their stroke-unaffected hand at any point. After each NF training run the participants were allowed to rest until they felt ready to continue onto the next run. Participants in the Sham group received identical instructions and tasks as participants in the Real group, however instead of seeing their own brain activity displayed in the NF bars, they saw a video of a previous participant’s feedback.

On each NF training day, before and after the NF training runs, participants completed Pre- and Post-Transfer runs. During these runs, participants were instructed to use movement strategies that they had found to be successful during the NF training runs, when they saw the instructions “Move Right” or “Move Left” (depending on which arm is stroke-affected) on the screen. However, unlike the NF training runs, they did not see the bars on the screen. During the transfer runs, participants received four 30-second movement blocks which were interspersed with five 30-second rest blocks during which the instructions changed to “Rest”. On the first NF training day, before having experienced any NF training runs, participants were instructed to practice movements with their stroke affected hand and received some suggested movements to try. After each NF training day, all participants completed a debriefing questionnaire in which they were asked about their perceived control over the two bars on a 5-point scale ranging from 1 (not in control) to 5 (fully in control).

### Motor performance measures

The primary outcome measure was performance on the JTT which was assessed on every testing day. The JTT is a timed assessment that measures performance of fine and gross hand function on 6 sub-tasks (handwriting task excluded) that simulate activities of daily living (e.g. simulated feeding). Participants were first familiarized with the task prior to the baseline assessment where they practiced the JTT sub-tasks until they reached stable performance. Timed data from the JTT was log transformed to improve normality as the residuals were significantly skewed as assessed by the Shapiro-Wilks test.

Secondary behavioral outcome measures included the ARAT and the UE-FM *(59)*. These measures were carried out on the baseline and follow-up testing days, to assess changes in stroke-related activity limitation and impairment respectively.

### Visuomotor squeeze task

Brain function during a controlled visuomotor squeeze task was assessed using fMRI at baseline and 1-week follow-up, and using EEG after each testing session (apart from 1-month follow-up). During the fMRI squeeze task, the participants held an MRI-compatible force transducer (BIOPAC, TSD121B-MRI) in their stroke affected hand whilst in the scanner. Before the start of the scan, the participant’s maximum squeeze force was recorded and a target force was determined (∼25% of max. force). During the scan, participants saw an empty grey bar on the screen. Periodically, a yellow line, which represented their target force, would appear across the grey bar, and participants were instructed to squeeze on the force transducer until a blue bar (which represented the current force exerted on the force transducer) reached the yellow line, at which point they should release. The frequency with which the yellow line appeared (i.e. frequency of squeezing) was set to a comfortable rate for each participant, but was typically between 0.25 and 0.35hz. Four 30-second squeezing blocks were interspersed with four 30-seconds rest blocks during which a fixation cross was presented on the screen.

The visuomotor squeeze task for EEG was similar to that described for fMRI. In each session, four movements blocks were performed, with rest breaks between. Each block consisted of 10, 5second trials, with 5.5-6.5second inter-trial interval. The number of squeezes performed in each trial was set individually for each participant, based on the frequency determined comfortable at baseline (range 1-4 squeezes). EEG data were collected using a 24 channel Ag/AgCl electrode EEG cap (Easycap, Germany) and Smarting mobile EEG Amplifier (mBrainTrain LL, Belgrade, Serbia) following standard scalp preparation techniques. Following pre-processing (see **Supplementary methods: EEG data analysis**), the event related desynchronization (ERD) was calculated for each trial for electrodes C3 and C4 (sensorimotor cortex). For the calculation of the ERD laterality index we used a procedure aligned to the fMRI data analysis, whereby values closer to 1 indicate greater lateralisation towards the affected hemisphere.

### MRI acquisition

MRI data was acquired on a 3 Tesla Siemens Magnetom Prisma MRI scanner (Siemens AG, Erlangen, Germany) using a 32-channel head coil. All task fMRI was acquired using multiband gradient echo-planer imaging. Additionally, during the baseline and 1-week follow-up scan a whole brain anatomical T1-weighted (MPRAGE) image was collected, as well as diffusion weighted imaging was acquired using a multi-shell, echo-planar imaging sequence (see **Supplementary methods: MRI acquisition**)

### MRI analysis

All fMRI data (NF, transfer and squeeze task runs) were pre-processed and co-registered using standard steps within FSL (see **Supplementary methods: MRI preprocessing**). Data from right-hand affected participants was mirror flipped. A first level GLM was carried out in FEAT for each fMRI run in order to compute task-based statistical parametric maps *(60)*. A double gamma-HRF convolved boxcar regressor, as well as its temporal derivative, was used to model the movement and rest blocks, and contrasts were set to Movement>Rest.

DWI data was preprocessed using FMRIB’s Diffusion Toolbox (see **Supplementary methods: MRI preprocessing**). The FA of the voxels in the CST ROIs was extracted and any outlier FA values (>3 IQR above median) were removed before averaging. Outlier values were usually restricted to the lesioned area, however in some participants these were also observed in perilesional tissue, near blood vessels or other abnormal tissues such as cavernomas. CST FA asymmetry was calculated as the difference in mean FA between the two CSTs (Unaffected - Affected)/(Unaffected+Affected), where positive values indicate lower FA/more damage in the stroke affected CST.

### Laterality and Success Index Calculation

In order to assess changes in laterality of brain activity, a LI was calculated based on the magnitude of the fMRI signal change in the ROIs used for NF training. Laterality indices are traditionally calculated as *(61)*:

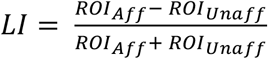

Where ROI_Aff_ and ROI_Unaff_ are fMRI activity in the affected and unaffected ROIs. This calculation yields a number between −1 and 1, with positive values indicating lateralization towards the affected hemisphere (higher values indicating stronger lateralization), and negative values indicating lateralization towards the unaffected hemisphere (lower values indication stronger lateralization). As it is possible that the average magnitude of activity in ROIs (as assessed using the average T-Stat value) is occasionally negative, which results in incorrect or meaningless LI values *(61)*, a threshold was applied to the activity in each ROI using the same approach as in work by Fernandez and colleagues *(62)*. In brief, for each ROI on each NF run, the mean maximum activity was calculated as the mean T-stat value of the top 5% of voxels showing the highest activity in the ROI. The threshold of activity that voxels needed to achieve for inclusion in the LI calculation was then set to 50% of this mean maximum activity. This ensured that only activity from the most active voxels was included in the LI calculation and that the average activity for each ROI was above 0. This approach has previously been validated as more robust and reliable than LI based on unthresholded signal intensity or voxel counts *(63)*.

A success index was calculated in order to capture variability in NF learning success. As in the current study NF learning appears to occur within (rather than across) NF training days, a NF slope success index was calculated over the runs on each day separately and then averaged. A regression slope was fit across the average LI over each of the NF runs within training days, and then the average slope was calculated over the three training days, resulting in a single NF success index.

### Statistical analysis

For statistical analysis of repeated measures data, linear mixed models (LMM) were used as implemented in RStudio (version 1.2.5) and the lme4 package *(64)*. LMMs allow for incomplete data, as well as greater flexibility in analysis, and better handling of repeated-measures dependencies *(65, 66)*. Random intercepts per participant were included in the random effects structures and the addition of random slopes per participant in the random effects structure was assessed using a likelihood ratio test *(67)*. There were no cases where the addition of random slopes significantly improved the model fit. P-values for fixed effects were derived using the Satterthwaite approximation for degrees of freedom, as implemented in the package lmerTest *(68)*. This approach has previously been shown to be less prone to type I errors, and is less sensitive to sample size than other methods *(69)*. Post-hoc comparisons were carried out using the emmeans package, with tukey method for multiple comparisons. We report unstandardized effect sizes (parameter estimates; *b*) whenever possible in line with general recommendations *(70)*, as there is currently no agreed upon way to calculate standard effect sizes for individual terms in LMMs.

For data with just a single follow-up (e.g. FA data) mixed ANCOVAs were carried out comparing the Real and the Sham group at follow-up, while accounting for baseline differences by including baseline as a covariate. Standard effect sizes are reported. The ANCOVA has previously been shown to be more sensitive in randomized studies compared to ANOVA of change from baseline *(71)*.

## Supporting information

CONSORT checklist

## Data Availability

Fully anonymized data from this study can be made available on request.

## Supplementary Materials

### Supplementary methods

**Fig. S1. Recruitment Flow chart**

**Fig. S2. Participants perceived control over NF bars**

**Fig. S3. Increased activity in the Putamen during initial NF learning**

**Fig. S4. No transfer effect on EEG visuomotor squeeze task**.

**Fig. S5. Baseline brain activity during visuomotor squeeze task is correlated with NF success**.

**Table S1. Average max. Tstat values for the affected and unaffected hemisphere**.

## Acknowledgments

The authors would like to thank Nele Demeyere and Grace Chiu for assistance with participant recruitment, Thomas Wassenaar for providing advice on statistical analysis and Chris Gallagher for technical support. This research was funded in whole, or in part, by the Wellcome Trust [Grant number 110027/Z/15/Z]. For the purpose of open access, the author has applied a CC BY public copyright licence to any Author Accepted Manuscript version arising from this submission.

## Data and materials availability

Fully anonymized data from this study can be made available on request.

