## Supplementary materials and methods for "Self-modulation of motor cortex activity after stroke alters behavior and corticospinal tract structure: a randomized controlled trial"

**Supplementary methods**

**Improvements over time on ARAT and UE-FM**

Two linear mixed models were carried out examining scores on the ARAT and UE-FM over all three timepoints (Baseline, 1-week and 1-month follow-up). Group and Day, as well as their interaction were included as fixed factors, and random intercepts were included in the random effects structure. A significant main effect of Day was observed on the ARAT (F(2,42.136)=3.323, p=0.046, linear trend: t(46.4)=2.055,p=0.045; *b*=1.86, SE=0.090), whilst the main effect of time fell short of significance on the UE-FM (F(2,42.225)=3.187, p=0.051). The main effect of Group and Group×Day interaction did not reach significance on either the ARAT or the UE-FM (all F< 1, p>0.5). This suggests both groups improved over time on these measures.

**Online analysis**

Functional images acquired during the NF training days were processed in real-time using Turbo-BrainVoyager 3.2 (Brain Innovation, Maastricht, Netherlands). This was achieved by transferring dicom images via a direct TCP/IP network link from the MRI console computer to the real-time computer. Images were then pre-processed in real-time, including head motion correction, spatial smoothing with a gaussian kernel of 3mm full-width-half-maximum (FWHM) and linear temporal drifts were removed from the data. A recursive GLM was carried out to compute T-Statistics for the contrast of Movement > Rest. The height of the bars (feedback) displayed to the participants was updated with each TR (933ms) and was calculated as follows:

FB = (CurrValue - Baseline) / Baseline × 100

Where CurrValue is the current BOLD signal in the ROI and Baseline is the average BOLD signal in the ROI during the last 20 TRs of the preceding Rest block. Only the last 20 TRs were included in the baseline calculation in order to exclude any possible BOLD decay from the previous movement block at the beginning of the Rest block. This resulted in a percent signal value that determined the height of the bar with respect to the maximum bar height which was set to double the PSC measured in the ROI during the functional localizer. Positive values being represented above the center line and negative values below it. The side on which the bars were displayed depended on which hand was stroke-affected, with the red bar always being displayed on the same side as the stroke-affected hand, and the blue bar on the same side as the stroke unaffected hand.

**EEG data acquisition**

Four blocks of task performance were performed per session, with rest breaks between. Each block consisted of 10, 5 s trials, with 5.5-6.5 s inter-trial interval. The number of squeezes performed in each trial was set individually for each participant in a familiarization session prior to the baseline assessments, based on the frequency determined comfortable for each person (range 1-4 squeezes). Participants viewed a laptop screen on a table in front of them and sat in a comfortable chair. Between trials a fixation cross appeared in the middle of the screen and participants were instructed to look at the cross and relax. When a yellow target line appeared on the screen participants were instructed to squeeze, and the height of a blue bar indicated the force applied to the force transducer in their affected hand. Participants were instructed to squeeze to reach the target and then release. At all times participants were instructed to keep their unaffected hand still and relaxed, to blink normally and to avoid looking around the room or clenching facial muscles. EEG data were collected using a 24 channel Ag/AgCl electrode EEG cap (Easycap, Germany; (www.easycap.de) and Smarting mobile EEG Amplifier (mBrainTrain LL, Belgrade, Serbia; <http://www.mbraintrain.com/>). Electrodes were positioned at 10–20 sites (Fp1, Fp2, Fz, F7, F8, FC1, FC2, Cz, C3, C4, T7, T8, CPz, CP1, CP2, CP5, CP6, TP9, TP10, Pz, P3, P4, O1, and O2). Reference and ground electrodes were placed at FCz and AFz sites respectively. Data were recorded with a sampling rate of 500 Hz (0–250 Hz pass-band) and transmitted wirelessly via Bluetooth to the Smarting Streamer App (resolution = 24 bit).

**EEG data analysis**

EEG data were analysed with EEGlab v14.1.2b. Data were cleaned using a well-established two-step procedure. Firstly, stereotypical artefacts were corrected, secondly epochs containing non-stereotypical artefacts were rejected. Specifically, the attenuation of stereotypical artefacts was done with independent component analysis (ICA). To optimize ICA decomposition quality, we performed several pre-processing steps before applying ICA. First, data were high-pass filtered (0.1 Hz, finite impulse response, filter order 8250) to remove low-frequency contributions (stationarity assumption). Second, data were low-pass filtered (40 Hz, finite impulse response, filter order 166) to remove high-frequency contributions (e.g. EMG activity, line noise) and subsequently data were down-sampled (250 Hz) to reduce computation time. Data containing non-stereotypical artefacts (e.g. gross head movements, swallowing) were removed. This was done by epoching the data into consecutive 1 s intervals and rejecting those containing non- stereotypical artefact (EEGLAB functions pop_ jointprob.m, pop_rejkurt.m, both SD = 4, rejected). This processing is considered an optimal trade-off for retaining much variance in the data but preventing the ICA algorithm being adversely affected by low- and high-frequency contributions as well as massive artefacts. Remaining data were submitted to extended infomax ICA to estimate the un-mixing weights of 24 independent components. Components representing stereotypical artefacts (e.g. eye blinks, eye movements, cardiac activity) were manually identified and removed from the raw data.

The artefact corrected raw data were then high-pass filtered at 8 Hz (finite impulse response, filter order 825) and subsequently low-pass filtered at 30 Hz (finite impulse response, filter order 220). EEG data were segmented (-2 s to 9 s relative to cue to begin task). The ERD% was calculated as follows:

ERD%(t) = (A(t)-R)/R x 100

where R is the power of a 1 s baseline interval (-1.5 to -0.5 s before the squeeze cue) and A is the power at time point t.

Outliers were detected using Grubbs test for each electrode and removed for that electrode only on a trial by trial basis. This was done to ensure maximal data retention given the relatively small number of trials completed each session.

For the calculation of the ERD laterality index we used a procedure aligned to the fMRI data analysis. For electrodes C3 and C4 the 95^th^ percentile of the ERD% over the task period was determined across all time points and all trials. The mean ERD% of the time points exceeding this threshold was calculated and then halved to determine the final threshold for inclusion. The mean ERD% of all time points exceeding this final threshold was determined (ERD%_MT_) and then used for calculation of the laterality index for a left hemisphere affected participant as follows:

LI = ( C3:ERD%_MT_ – C4:ERD%_MT_ ) / ( C3:ERD%_MT_ + C4:ERD%_MT_)^[1]^

whereby values from -1 to 1 represent ERD lateralised to the unaffected hemisphere through to ERD lateralised to the affected hemisphere. For a right hemisphere affected participant the placement of C3 vs C4 is swapped in the formula.

**MRI acquisition**

FMRI during NF training, as well as for the fMRI squeeze task, was acquired using multiband gradient echo-planer imaging (*(72)*; TR=933ms, TE=33.4ms, flip angle=60°, slice thickness = 2mm, in-plane resolution = 2×2, 72 slices, multiband factor = 6). Additionally, a high-contrast (single-band) whole brain EPI image was acquired for registration purposes, and a dual echo fieldmap for fieldmap unwarping (TR = 482ms, TE1 = 4.92ms, TE2 = 7.38ms, flip angle=46°,slice thickness = 2mm, 49 slices).

Whole brain T1-weighted (MPRAGE) images were acquired at baseline and 1-week follow-up with 1mm resolution (TR = 1900ms, TE = 3.97ms, flip angle = 8°, TI = 904ms, slices = 192). Additionally, diffusion weighted imaging (DWI) was acquired at baseline and 1-week follow-up using a multi-shell, echo-planar imaging sequence (TR = 2483ms, TE = 78.2ms, voxel-size = 1.75 × 1.75 × 1.75 mm^3^, 76 slices, multiband factor = 4) with 11 non diffusion-weighted images (*b*= ~0s/mm) and 60 diffusion-weighted images acquired with a b-value of 1500 s/mm and 60 with a b-value of 2500 s/mm. This data was acquired in both the A>P and the P>A phase encoding direction.

**MRI pre-processing**

FMRI data was preprocessed using standard preprocessing steps including motion correction, brain extraction, fieldmap-based EPI distortion correction using FUGUE, spatial smoothing using a 5mm FWHM Gaussian kernel and high pass temporal filtering with a cut-off of 90 seconds. Co-registration was achieved by first using linear registration (FLIRT; *(73)* to register the multiband functional data to a single volume, high-contrast (single-band) EPI image. Next, the high-contrast EPI image was linearly registered to the T1-weighted image using boundary-based registration (6 degrees of freedom;*(74)*. For voxelwise group analysis, nonlinear registration (FNIRT) into MNI space was carried out using lesion masks to exclude voxels in the lesioned area. For NF runs, intermediate fixed effects analysis steps were carried out in order to average run activity across NF days (average of each run over days). Intermediate fixed effects analysis was carried out to create difference maps between days and runs (e.g. Baseline vs Follow-up, Average Run1 vs Average Run3 etc.). Group level analysis was carried out using FMRIB’s Local Analysis of Mixed Effects (FLAME) to compare difference maps between the Real and the Sham groups *(75)*. Group Z-statistic images were thresholded using clusters determined by Z > 3.1 and a family-wise corrected cluster significance threshold of p<0.05 was applied.

DWI data was preprocessed using FMRIB’s Diffusion Toolbox (FDT). Pre-processing steps included distortion correction using *topup* and eddy-current correction using *eddy.* FA maps were then created by fitting a diffusion kurtosis tensor at each voxel using *dtifit,* and were brain extracted*.* Diffusion kurtosis imaging (DKI) allows quantification of non-gaussian diffusion and has previously been shown to produce more sensitive and accurate tissue quantification *(76)*. The halfway linear registration between the DKI-derived FA maps from the baseline and follow-up session was calculated, and the FA maps were linearly-registered to this halfway space and averaged to create a midspace image. The midspace image was then non-linearly registered to the FMRIB FA template in MNI space using ANTs (Advanced Normalization Tools; *(77)*). Lesion masks were used to exclude lesion areas from the registration. CST ROIs were delineated in standard space from the JHU White-Matter Tractography atlas. These ROIs were then registered into the native participant- and time-point specific space using the transformations generated by ANTs. If the lesioned area encroached upon one of these ROIs, the overlapping voxels were removed.

**Missing Data**

During the NF training sessions the following data was missing: Due to excessive motion, defined as average absolute motion >2 standard deviation above group mean motion, 7 individual NF runs (2 in the Real group from the same participant, 5 in the Sham group with three runs from the same participant and two from different participants) were removed. One run was also removed from the Real group due to a technical issue with the NF software. Additionally, two transfer runs were removed from the Sham group due to excessive motion in separate participants, one transfer run was not collected due to the participant stopping the scan early, and all of the transfer data for one participant in the Real group was removed as 5/6 transfer runs had excessive motion.

For the Baseline and 1-week follow-up sessions, two participants (one from each group) did not complete the follow-up scan (one due to feeling unwell, one due to the scanner being out of use). Both of these participants still completed the follow-up motor assessments. Additionally, DWI data was missing from one participant in the Real group who requested to stop the scan early. For the fMRI visuomotor squeeze task, in addition to the two participants who did not complete the follow-up scan, two participants in the Sham group had missing data, one due to hardware issues with the force transducer and one due to excessive motion during the baseline scan. For the EEG visuomotor squeeze task, data from two participants was removed entirely due to persistent problems with the recorded signal. In four further participants data from one or two testing sessions was missing (corrupted or unreadable data), however their remaining data was included in analyses.

Finally, six participants (five in the Real group and one in the Sham group) did not complete the one-month follow-up for varying reasons (see recruitment flow chart **Supplementary Figure S1**).

**Suggested strategies**

The following strategies were suggested and demonstrated to the participants:

- Opening and closing your affected hand
- Moving individual fingers of your affected hand
- Moving your affected hand at the wrist
- Tapping the fingers of your affected hand onto your thumb

Additional suggested strategies:

- Focusing less on your unaffected hand
- Focusing more on your affected hand
- Relaxing your unaffected hand while moving your affected hand
- Imagining movements of the affected hand
- Combining different movements

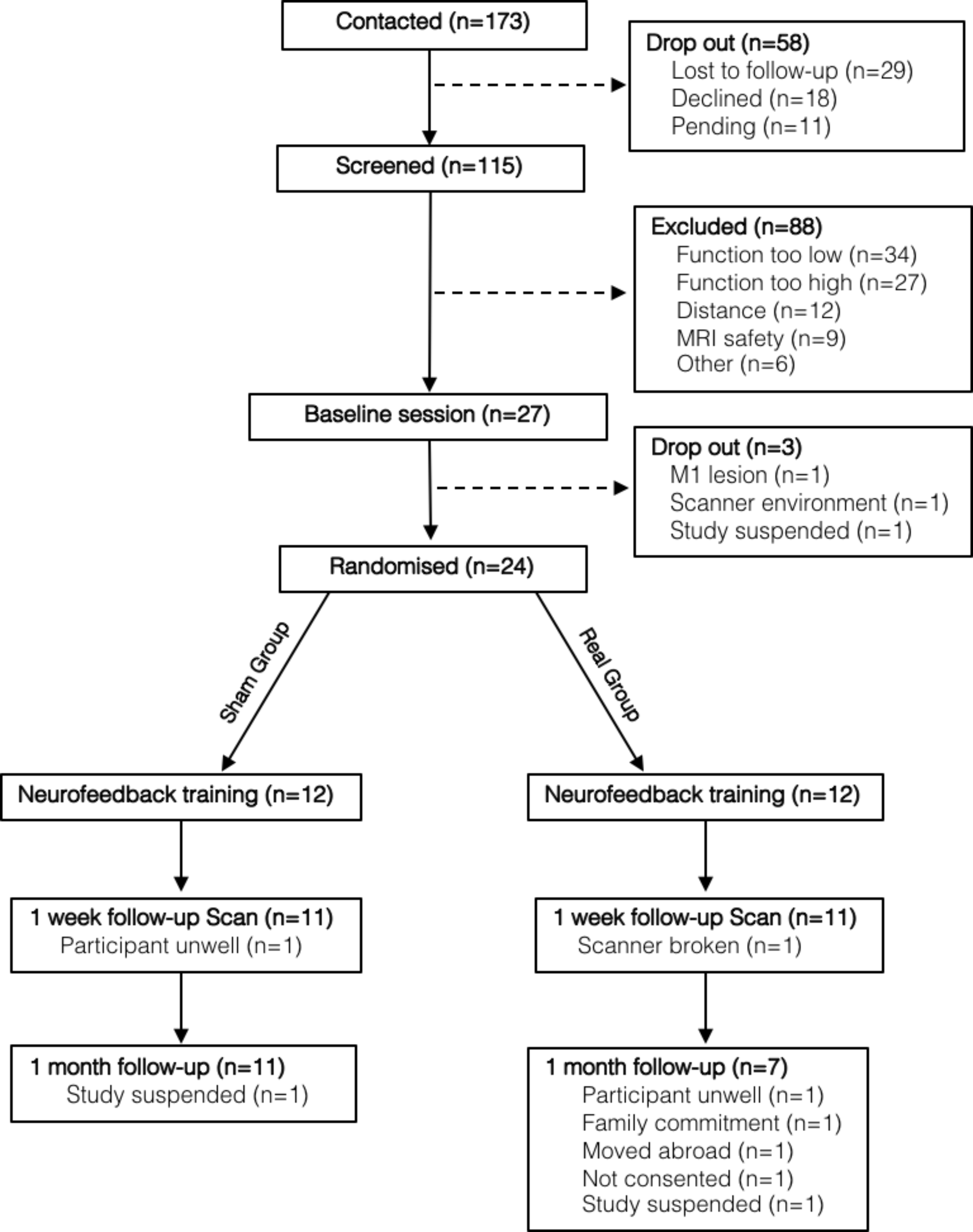

**Supplementary figure S1: Recruitment flow chart.** Number of patients at each stage indicated by *n*, as well as the reason for drop out or exclusion. The participant from the sham group who was unable to undergo the 1 week follow up scan was still able to undergo behavioral assessments. Note the study was halted prematurely due to restrictions from the coronavirus pandemic, resulting in 11 potential participants that we were unable to assess for eligibility.

**Supplementary figure S2: Participants perceived control over NF bars.** Participants rated their perceived control over the NF bars after each session on a scale from 1–5 (not in control – fully in control). Average ratings over the three training days is displayed, with the bar representing the median response and individual participants’ data shown as black dots (Real) or diamonds (Sham). Participants generally reported more control over the red bar reflecting activity in the stroke affected hemisphere than over the blue bar, reflecting unaffected hemisphere activity. This was confirmed using a related-samples Wilcoxon signed ranks test which showed significantly lower perceived control for the blue bar than the red (Z=-3.17, p=0.002, *d*=4.539). Mann-Whitney U test revealed no significant differences between the two groups for the two bars (Red bar: U = 51, Z=-1.317, p=0.188, *d* = 0.511; Blue bar: U = 49, Z=-1.389, p=0.165, *d* = 0.563).

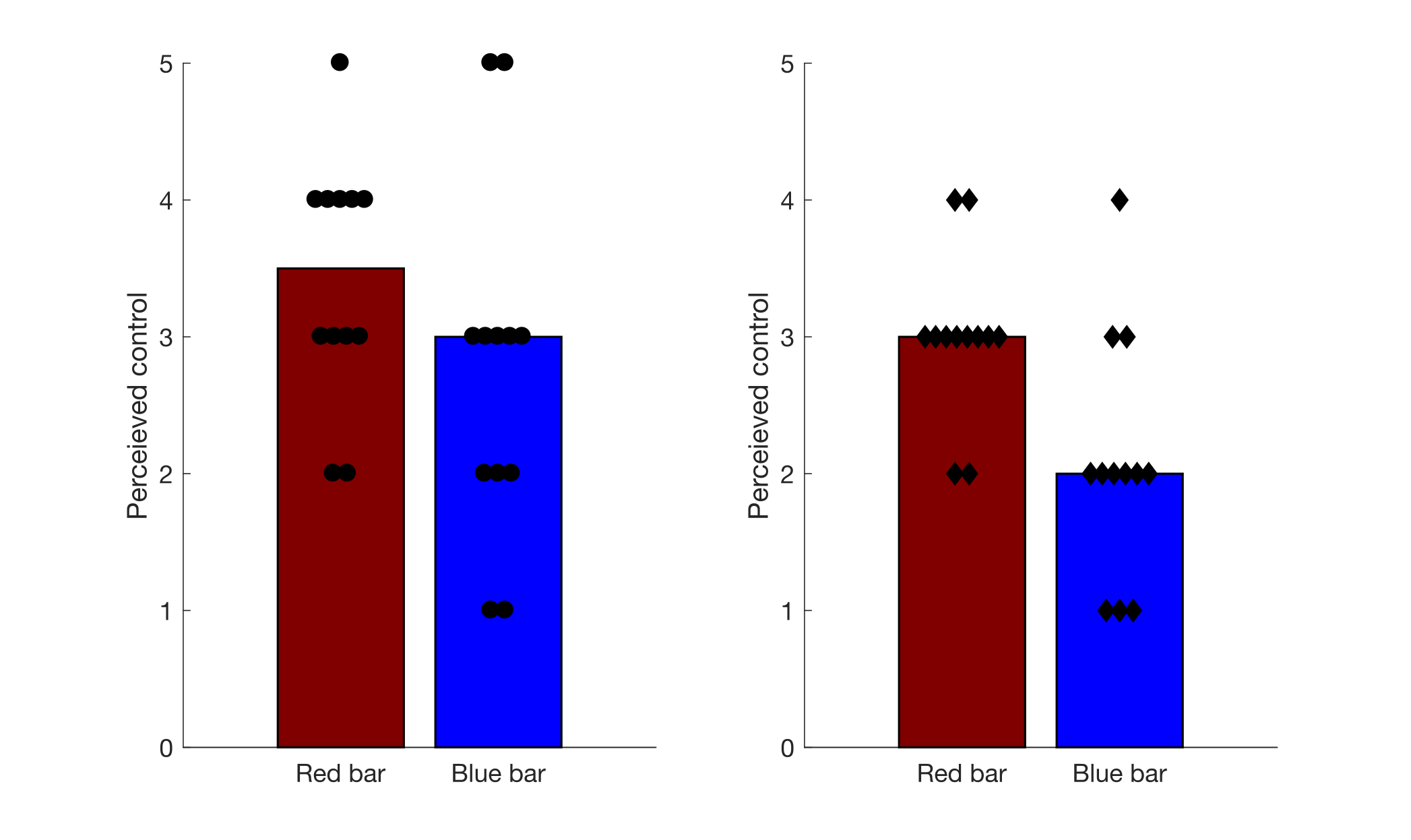

Real

Sham

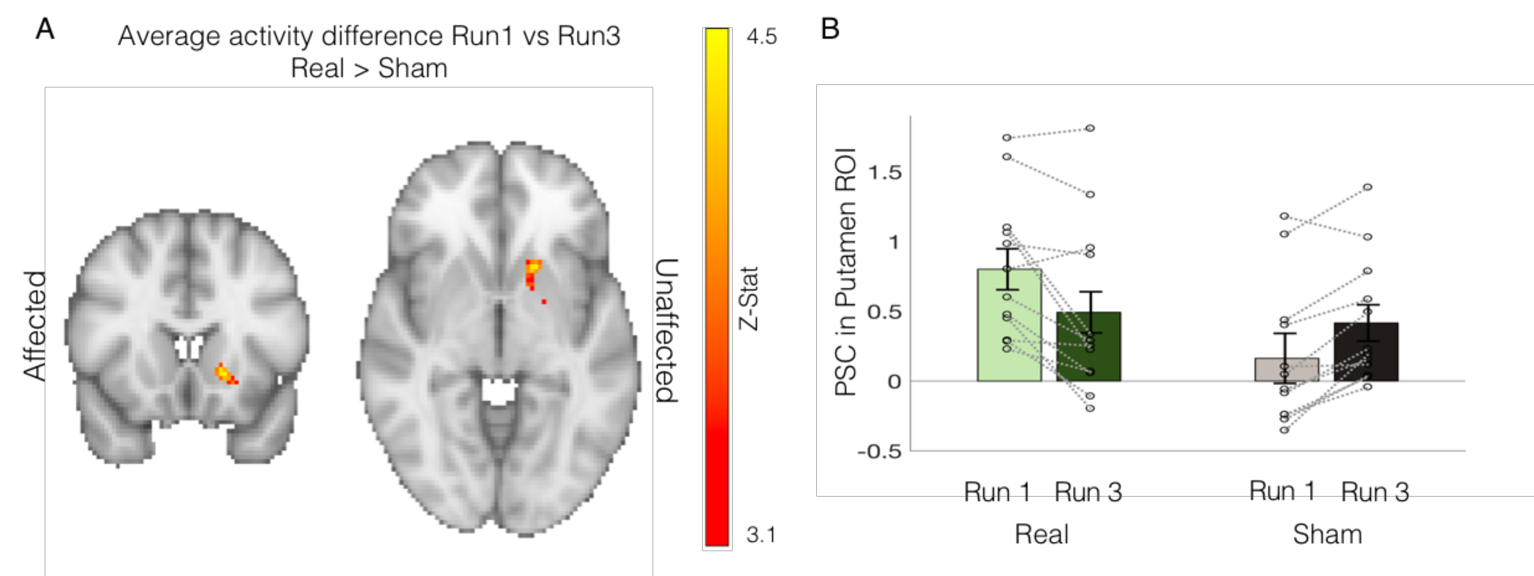
**Supplementary figure S3: Increased activity in the Putamen during initial NF learning. (A)** A significant cluster was found in the putamen of the unaffected hemisphere where change in activity between the first and the last NF runs on each day (Average Run1 > Average Run3) was greater in the Real group than the Sham (Real>Sham, voxelwise GLM, p<0.05, corrected). **(B)** For visualization purposes, the mean PSC of the significant clusters is plotted on the right (Real = Green; Sham = Grey), as well as the data from individual participants (represented by open circles). Error bars represent s.e.m.

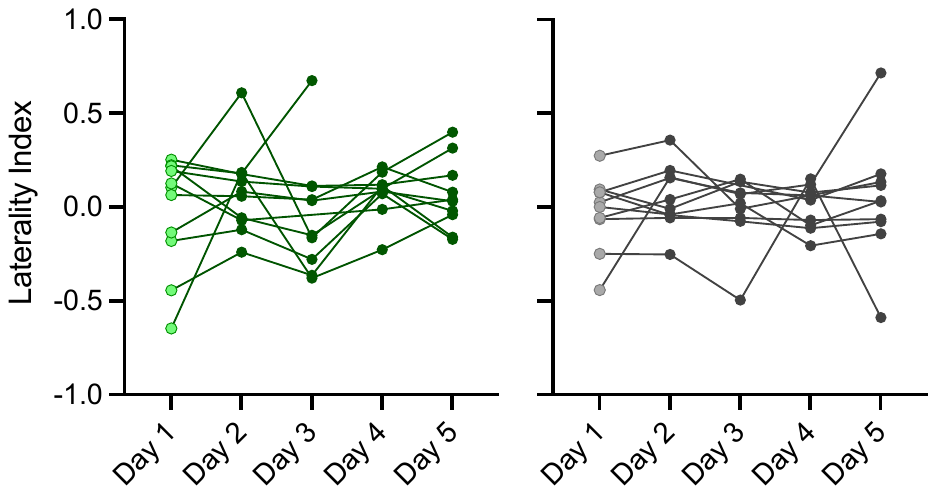

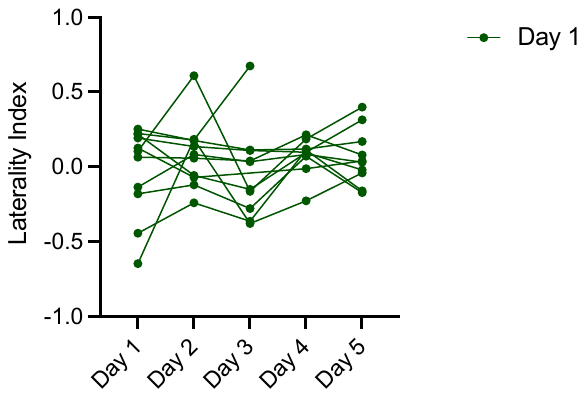

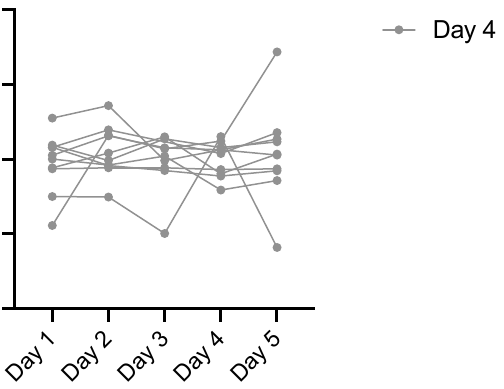

Real

Sham

**Supplementary figure S4: No transfer effect on EEG visuomotor squeeze task.** For the EEG squeeze task, LI was calculated based on the event related desynchronization (ERD) obtained from electrodes C3 and C4. LI is displayed for each participant on each day for the Real (green/right) and the Sham (grey/left) group. A LMM was carried out on the LI with Group and Day as fixed effects, baseline LI as a covariate and random intercepts in the random effects structure. There was no effect of Group (F(1,48)=0.003, p=0.958) or Day (F(1,58)=0.119, p=0.731), and no interaction (F(1,58)=0.303, p=0.584. There was also no correlation between follow-up LI and NF success index in the Real group (rho=0.273, p=0.446, n=10, two-tailed). This suggests that the laterality of motor activity on this task did not change significantly over the course of the NF training.

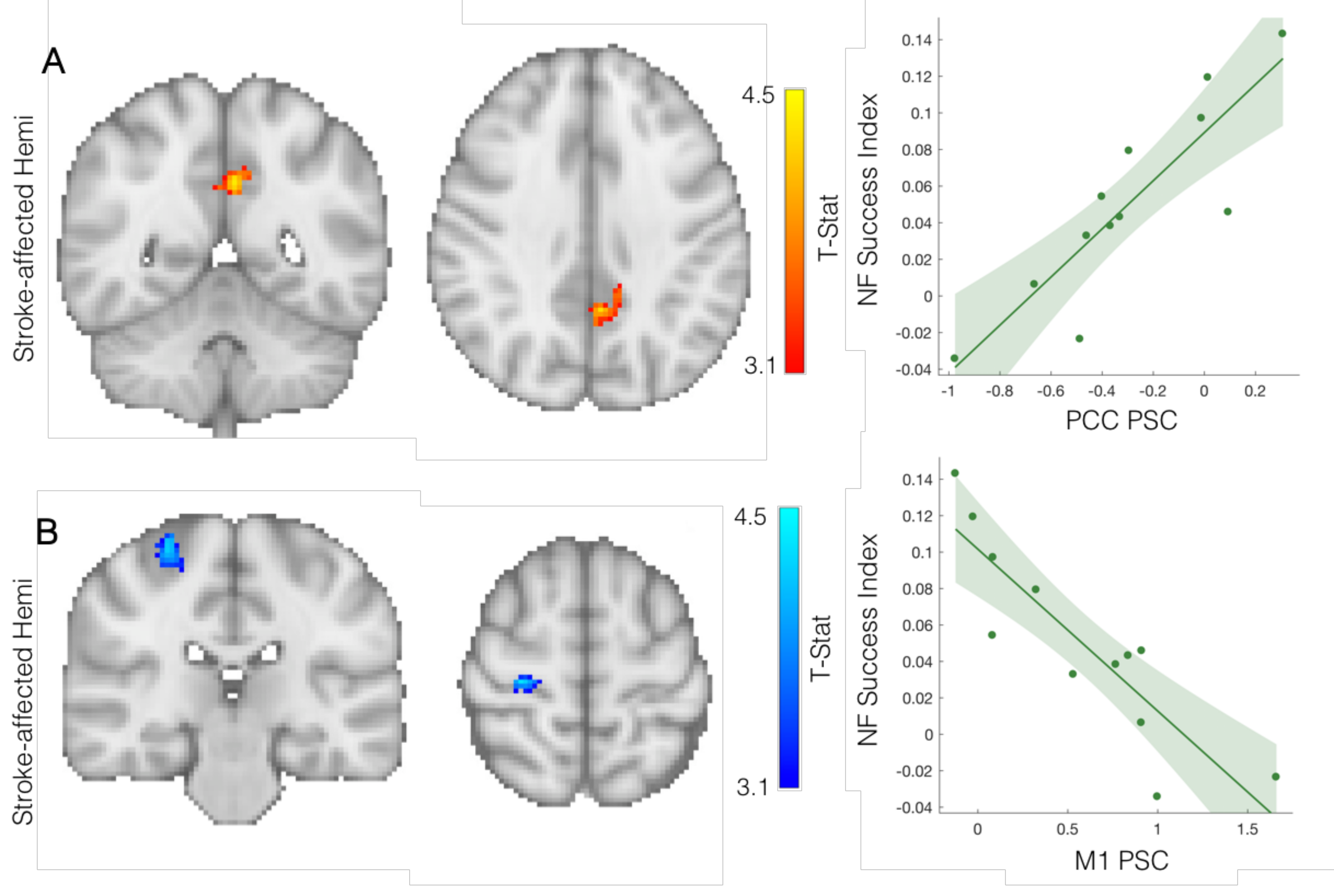

**Supplementary figure S5: Baseline brain activity during visuomotor squeeze task is correlated with NF success. (A)** Voxel-wise analysis revealed a cluster in the Posterior Cingulate Cortex (PCC; 145 voxels, p=0.001, MNI coordinates of max. zstat: -4 -50 34 ) where a significant positive correlation (displayed in red/yellow) between activity and the NF success index was found. Scatter plot for visualisation purposes shows PSC of voxels within the significant cluster in relation to the SI. **(B)** Additionally a significant cluster was also found in the stroke-affected motor cortex (96 voxels, p=0.015, MNI coordinates of max. zstat: 28 -24 60) where a significant negative correlation (displayed in blue) was found between brain activity at baseline and NF success index. Again the scatter plot shows the PSC of the significant voxels plotted against the SI for visualisation purposes only. Shaded area corresponds to 95% confidence interval.

|  |  | **Affected** | | **Unaffected** | |
| --- | --- | --- | --- | --- | --- |
|  |  | Real | Sham | Real | Sham |
|  | Run 1 | 8.311 (1.234) | 8.355 (1.334) | 5.227 (0.696) | 4.292 (0.775) |
| Day 1 | Run 2 | 9.141 (1.792) | 7.573 (1.165) | 5.880 (1.338) | 4.912 (0.749) |
|  | Run 3 | 9.424 (1.787) | 7.239 (1.374) | 5.395 (1.005) | 4.364 (0.873) |
|  | Run 1 | 7.564 (1.096) | 7.889 (1.182) | 4.734 (0.790) | 5.371 (0.783) |
| Day 2 | Run 2 | 8.337 (1.173) | 7.732 (1.014) | 4.750 (0.783) | 5.359 (0.796) |
|  | Run 3 | 8.166 (1.333) | 8.048 (1.469) | 4.504 (0.870) | 4.906 (1.067) |
|  | Run 1 | 8.068 (1.264) | 8.937 (1.680) | 5.095 (0.870) | 5.242 (1.054) |
| Day 3 | Run 2 | 8.369 (1.347) | 7.388 (1.174) | 5.451 (1.004) | 4.613 (0.751) |
|  | Run 3 | 8.305 (1.456) | 8.660 (1.812) | 4.959 (0.971) | 5.232 (0.827) |

**Table S1: Average max. Tstat values for the affected and unaffected hemisphere.** Mean max. Tstat value used for computing the LI is shown per hemisphere for each run on each training day (s.e.m is shown in brackets). To examine whether LI changes within training days were driven more by activity changes in the affected or unaffected M1, activity in each ROI was analyzed separately using LMMs. There were no significant main effects or interaction in either the affected or the unaffected M1 ROI (all F <2.75, all p >0.07).

1. Results are the same if laterality is computed as: C3-C4.
